# A Phase Ib/II multi-arm, dose finding and expansion study of a novel thymidylate synthase inhibitor with immune modulating properties, NUC-3373, in combination with pembrolizumab or docetaxel in patients with advanced solid tumors (NuTide:303)

**DOI:** 10.1101/2024.11.07.24316829

**Authors:** Gary Middleton, James Spicer, Burak Aktas, Habana Dinizulu, Richard H Wilson, David J Harrison, Elisabeth Oelmann, Jeff Bloss, Fiona Thistlethwaite

## Abstract

**Background:** NUC-3373 is a novel thymidylate synthase (TS) inhibitor designed to directly deliver the active anti-cancer metabolite fluorodeoxyuridine-monophosphate (FUDR-MP or FdUMP) intracellularly. In addition to being a potent TS inhibitor, NUC-3373 has also been shown to cause DNA damage and promote the release of damage-associated molecular patterns. These diverse mechanisms position NUC-3373 as a potentially effective combination partner for several other anti-cancer agents.

**Methods:** NuTide:303 was a Phase Ib/II open label, multi-arm, parallel cohort dose-finding and expansion study designed to evaluate optimal combination partners for NUC-3373. Module 1 planned to enroll up to 12 evaluable patients with advanced/metastatic solid tumors to determine the recommended dose for NUC-3373 in combination with leucovorin (LV) and pembrolizumab. Module 2 planned to enroll approximately 6-12 evaluable patients with advanced/metastatic non-small cell lung cancer (of any histology) or pleural mesothelioma to determine the recommended dose for NUC-3373 in combination with LV and docetaxel. A 3+3 dose escalation design was used in both modules, with safety parameters continually assessed and tumor assessments conducted at screening and every 8 weeks from C1D1 until progression. The study was terminated early and the Phase II part was not initiated; therefore, only results of the Phase Ib part of the study are presented.

**Results:** Fifteen patients were enrolled in Module 1, of whom 13 received study treatment and were included in the safety population. Nine patients were evaluable for anti-tumor activity. No DLTs were observed in these heavily pre-treated PD-(L)1 experienced patients with a variety of different tumor types. An objective response rate of 22% and a disease control rate of 67% were reported. NUC-3373 + LV + pembrolizumab was well tolerated, the most common treatment-related adverse events (AEs) were nausea, vomiting, diarrhea, and fatigue and the majority of events were Grade 1 or 2. Four patients were enrolled in Module 2, all of whom received study treatment. Three evaluable for anti-tumor activity. Two patients treated at the first dose level achieved stable disease and completed >6 months of treatment. The most common treatment-related AE was fatigue (n=4) and the majority of events were Grade 1 or 2.

**Conclusions:** The results from the Phase Ib part of NuTide:303 suggest that NUC-3373 may be an effective combination partner for pembrolizumab in a variety of advanced solid tumors. The probability of being able to escalate to typical efficacious doses of docetaxel used in combination is low due to overlapping toxicity. The use of a different taxane in this combination is currently being considered.

## Introduction

NUC-3373 is a novel thymidylate synthase (TS) inhibitor that was specifically designed to overcome the key limitations and pharmacologic challenges associated with fluoropyrimidines and other TS inhibitors [1, 2]. TS is a crucial enzyme in the *de novo* synthesis of deoxythymidine monophosphate (dTMP), a nucleotide essential for DNA synthesis and repair [3]. TS is often upregulated in rapidly dividing cancer cells, making it a critical target for anti-cancer agents. There are several widely used anti-cancer agents that inhibit TS, including 5-fluorouracil (5-FU), capecitabine and pemetrexed; however, their clinical utility is limited by off-target toxicity, inefficient activation pathways, and poor pharmacokinetic (PK) profiles [4-7].

NUC-3373 was designed to directly deliver the active anti-cancer metabolite fluorodeoxyuridine-monophosphate (FUDR-MP or FdUMP) intracellularly, bypassing the complex and inefficient activation pathway that is required for other fluoropyrimidines. NUC-3373 is also resistant to enzymatic breakdown by dihydropyrimidine dehydrogenase (DPD), reducing the generation of the toxic by-product α-fluoro-β-alanine (FBAL) and offering a potential treatment option to patients with DPD deficiencies. We have shown that in addition to being a potent TS inhibitor, NUC-3373 is an effective DNA-damaging agent [8] and promotes release of damage-associated molecular patterns (DAMPs) [9, 10]. These diverse mechanisms position NUC-3373 as a potentially effective combination partner for several other anti-cancer agents, including checkpoint inhibitors.

NuTide:303 was a Phase Ib/II open label, multi-arm, parallel cohort dose-finding and expansion study designed to evaluate optimal combination partners for NUC-3373 in patients with advanced solid tumors [11]. Two modules were opened. Module 1 was designed to investigate NUC-3373 in combination with leucovorin (LV) and pembrolizumab for the treatment of patients with advanced solid tumors. The rationale for this combination is based on data from nonclinical studies showing that NUC-3373 may have the potential to modulate the tumor microenvironment (TME) and prime an anti-tumor immune response. In colorectal cancer (CRC) cell lines, NUC-3373 was shown to cause DNA damage and promote the release of DAMPs, including high-mobility-group box 1 (HMGB1), calreticulin and ATP [9, 10]. DAMPs are known to enhance cancer cell recognition by the immune system, promoting recognition and destruction by inflammatory and immune mechanisms. In co-culture experiments, NUC-3373 treatment caused an increase in the surface expression of PD-L1 on CRC and non-small cell lung cancer (NSCLC) cells, indicating that NUC-3373 may enhance the anti-cancer potential of immune checkpoint inhibitors targeting the PD-1/PD L1 axis [12, 13]. In support of this, CRC and NSCLC cells pre-treated with NUC-3373 and co-cultured with peripheral blood mononuclear cells (PBMCs) displayed a reduction in cell viability compared to monocultured CRC or NSCLC cells. Furthermore, NUC-3373 was shown to enhance cell death in CRC and NSCLC cells co-cultured with PBMCs, with NUC-3373 potentiating the activity of the PD-1 inhibitors nivolumab (in CRC cells) and pembrolizumab (in NSCLC cells) [12, 13]. Thus, NUC-3373 is an attractive combination partner for PD-(L)1 inhibitors.

Module 2 was designed to investigate NUC-3373 in combination with LV and docetaxel for the treatment of patients with NSCLC or pleural mesothelioma. The rationale for this module is based on data from nonclinical studies showing that NUC-3373 is a more potent TS inhibitor than either 5-FU or pemetrexed in NSCLC cells. In a set of *in vitro* studies in four human NSCLC cell lines, NUC-3373 showed cytotoxicity across all cell lines with half-maximal inhibitory concentration (IC50) values in the low µM range. Consistent with existing literature, the squamous cell carcinoma cells lines (Cx140 and Nx002) had higher TS expression levels than the adenocarcinoma cell lines (A549 and Calu3) [14]. However, treatment with NUC-3373 promoted TS inhibition in all four cell lines, showing that NUC-3373 is able to inhibit TS regardless of basal TS expression levels and cell histology [14]. These results suggest that NUC-3373 may have activity where pemetrexed has previously failed in lung cancers with squamous cell differentiation.

The nonclinical findings have been substantiated in patients treated with NUC-3373 as a monotherapy and in combination with other agents in clinical studies [15-19]. NUC-3373 has been well-tolerated and has shown a favorable PK profile.

These data demonstrate the potential for NUC-3373 to offer an effective treatment option and combination partner in several tumor indications.

The NuTide:303 clinical study was terminated early due to refinement of the pipeline strategy not due to any adverse safety signal. Phase II part of the study was not initiated. Here we describe the results from the Phase Ib part of Modules 1 and 2.

## Methods

### Study Design and Objectives

NuTide:303 was a Phase Ib/II open label, multi-arm, parallel cohort dose-finding and expansion study designed to evaluate the safety, efficacy and PK of NUC-3373 in combination with other agents for the treatment of patients with different tumor types (EudraCT 2022-000722-14; NCT05714553). This study was modular, with each module evaluating a different NUC-3373 combination, and consisted of a dose-validation phase (Phase Ib) and a potential dose-expansion phase (Phase II). Approximately 6-20 evaluable patients were planned to be enrolled in the Phase Ib part of each module to determine safety, tolerability, and preliminary efficacy of NUC-3373 in combination with other agents. Following review of the Phase Ib data, each module may then have moved into Phase II to enable a further assessment of safety and efficacy in approximately 20-40 patients.

In Module 1, up to 12 evaluable patients were planned to be enrolled in Phase Ib to determine the recommended dose for NUC-3373 + LV in combination with pembrolizumab. In Phase II, a further evaluation of the combination was planned in exploratory, signal-seeking cohorts of approximately 20-40 patients in 2-4 selected tumor types that have shown encouraging signs of activity (clinical or translational) during Phase Ib. Phase II portion of study was not initiated.

In Module 2, approximately 6-12 evaluable patients were planned to be enrolled in Phase Ib to determine the recommended dose for NUC-3373 + LV in combination with docetaxel. In Phase II, a further evaluation of the combination using a Simon’s two-stage design was planned [20]. Phase II portion of study was not initiated.

The study was conducted at 3 clinical sites in the United Kingdom and was performed in accordance with the Declaration of Helsinki and the principles of Good Clinical Practice. The protocol was approved by the relevant Ethics Committees and all patients provided written informed consent before undergoing any study procedures.

### Patients

#### Module 1

Patients with advanced/metastatic solid tumors who had progressed on ≤2 prior therapies for metastatic disease, that may have included 1 prior immunotherapy-containing regimen (either monotherapy or in combination with chemotherapy) or who had not progressed but where addition of NUC-3373 + LV to standard pembrolizumab monotherapy may have been appropriate (*i*.*e*., patients who had exhausted all other treatment options for advanced disease) were eligible. Patients must have had tumor types for which pembrolizumab is approved and is a suitable treatment option (e.g., melanoma, NSCLC, urothelial carcinoma, esophageal carcinoma, renal cell carcinoma, head and neck squamous cell carcinoma, cutaneous squamous cell carcinoma, MSI high CRC, gastric cancer, triple negative breast cancer, classical Hodgkin lymphoma, and endometrial carcinoma). Patients must also have had radiologically measurable disease, Eastern Cooperative Oncology Group (ECOG) performance status 0-1, and adequate hematologic, renal and hepatic function. Patients with known neutralizing antibodies against checkpoint inhibitors or who had discontinued prior therapy due to toxicity attributed to checkpoint inhibitors were excluded.

#### Module 2

Patients with advanced/metastatic NSCLC (of any histology) or pleural mesothelioma who had progressed on, or were unable to tolerate, 1 or 2 prior lines of cytotoxic chemotherapy-containing regimens for advanced/metastatic disease (not including neoadjuvant or adjuvant therapy) were eligible. Additional prior lines of treatment with targeted agents or immunotherapy were allowed as long as they were not given in combination with cytotoxic chemotherapy. Patients must also have had radiologically measurable disease, ECOG performance status 0-1, and adequate hematologic, renal and hepatic function. Patients who had received prior treatment with docetaxel for advanced/metastatic disease were excluded.

### Treatment

#### Module 1

An initial cohort of 3 evaluable patients were to receive NUC-3373 at a dose of 1875 mg/m^2^ plus LV (400 mg/m^2^) on Days 1, 8 and 15 in combination with pembrolizumab (200 mg) on Day 1 of 21-day cycles. If this dose was tolerated (i.e., no DLTs in the first 3 patients), a further 3 patients were to be enrolled at the same dose level. Following this, the cohort could have been expanded up to a total of 12 evaluable patients.

#### Module 2

An initial cohort of 3 evaluable patients were to receive NUC-3373 at a dose of 750 mg/m^2^ plus LV (400 mg/m^2^) on Days 1 and 22 and docetaxel (55 mg/m^2^) on Day 8 of 28-day cycles. If this dose was tolerated, dose escalation was to proceed in a modified 3+3 design. The dose and schedule for docetaxel was slightly modified from the standard regimen used in advanced NSCLC, in which docetaxel is typically administered every 3 weeks at a dose of 75 mg/m^2^, to a 4-weekly administration schedule. This modification was for safety reasons to avoid concomitant treatment with a new agent (NUC-3373) during the post-docetaxel white blood cell nadir. Following treatment of 4 patients at the first dose level, dosing was stopped due to toxicity challenges with docetaxel, which required dose reductions to 35 mg/m^2^ in 1 patient from Cycle 3 onwards.

### Clinical Assessment

All patients who received at least one dose of study treatment were included in the safety analysis population. Safety parameters were continually assessed. Treatment-emergent adverse events (TEAEs) were graded according to the National Cancer Institute Common Terminology Criteria for Adverse Events. Patients were defined as evaluable for DLT assessment if they had received at least 75% of the intended dose of each agent during Cycle 1 (Module 1) or Cycles 1 and 2 (Module 2) or experienced a DLT. The definition of DLT is provided in the Supplementary Data.

Tumor response assessments were performed for the evaluable population, defined as patients with measurable disease at baseline who received at least two cycles of treatment, received at least 75% of planned treatment over the two cycles, and had a post-treatment objective disease assessment. Computed tomography (CT)/magnetic resonance imaging (MRI) based tumor assessments were conducted at screening and every 8 weeks from C1D1 until progression. Responses from all available post-baseline scans from evaluable patients were used for derivation of the best overall response (BOR) and disease control rate (DCR). A response of stable disease was only considered if the scan was taken at least 7 weeks after the baseline scan. The highest reduction of the sum of diameters of target lesions was selected and reported for each patient with available post-baseline scan. This was expressed as a percentage change from baseline. In Module 1, the primary efficacy endpoint (ORR) was assessed using iRECIST criteria and the secondary efficacy response endpoints were assessed using RECIST v1.1 criteria. In Module 2, all efficacy response endpoints were assessed using RECIST v1.1 criteria. Responses (CRs and PRs) had to be confirmed by repeated measurement at least 28 days and not more than 42 days after initial documentation. As per iRECIST, assessment of disease progression also required confirmation of PD in Module 1. Patients were to be treated and followed-up post-initial progression (termed unconfirmed PD or iUPD) until subsequent disease progression (termed confirmed PD or iCPD) was confirmed.

### Statistical Analyses

Since this was an exploratory study focused on tolerability, translational data, and signal-seeking activities, descriptive statistics were used.

#### Module 1

In Phase Ib, up to approximately 12 evaluable patients in total were to be enrolled to determine the tolerability of NUC-3373 in combination with pembrolizumab.

#### Module 2

In Phase Ib, approximately 6-12 evaluable patients were to be enrolled in a 3+3 dose escalation design to determine the recommended dose for NUC-3373 in combination with docetaxel.

The data presented have a cut-off date of 30 May 2025, at which time the Sponsor terminated the NuTide:303 study early; therefore, the Phase II part of the study was not initiated. This was primarily due to refinement of the pipeline strategy and not due to an adverse safety signal. At the end of the study, one patient in Module 1 continued to receive treatment with NUC-3373 + pembrolizumab in a post-trial capacity. The patient remains in a durable partial response (81% reduction) at manuscript submission (personal communication, FT, The Christie NHS Foundation Trust, August 2025).

## Results

### Patient Characteristics

Between March 2023 and July 2024, 19 patients were enrolled across 3 centers in the UK (Table 1).

**Table 1.** Patient baseline characteristics.

|  | Module 1 (n=13)* | Module 2 (n=4) |
| --- | --- | --- |
| Median age (range) | 68 (49-75) | 67 (60-75) |
| Sex, m/f | 11/2 | 2/2 |
| ECOG PS, 0/1 | 4/9 | 1/3 |
| **Target lesions** |  |  |
| 1 | 5 | 1 |
| 2-3 | 6 | 1 |
| ≥4 | 2 | 2 |
| Liver, yes/no | 3/10 | 0/4 |
| Lung, yes/no | 8/5 | 2/2 |
| Median prior lines (range)# | 1 (0-2) | 2 (1-3) |
| Abbreviations: ECOG PS=Eastern Cooperative Oncology Group performance status |  |  |
| *2 patients in Module 1 were not safety evaluable as they no longer met eligibility requirements at C1D1 |  |  |
| # for metastatic disease |  |  |

**Table 2.**
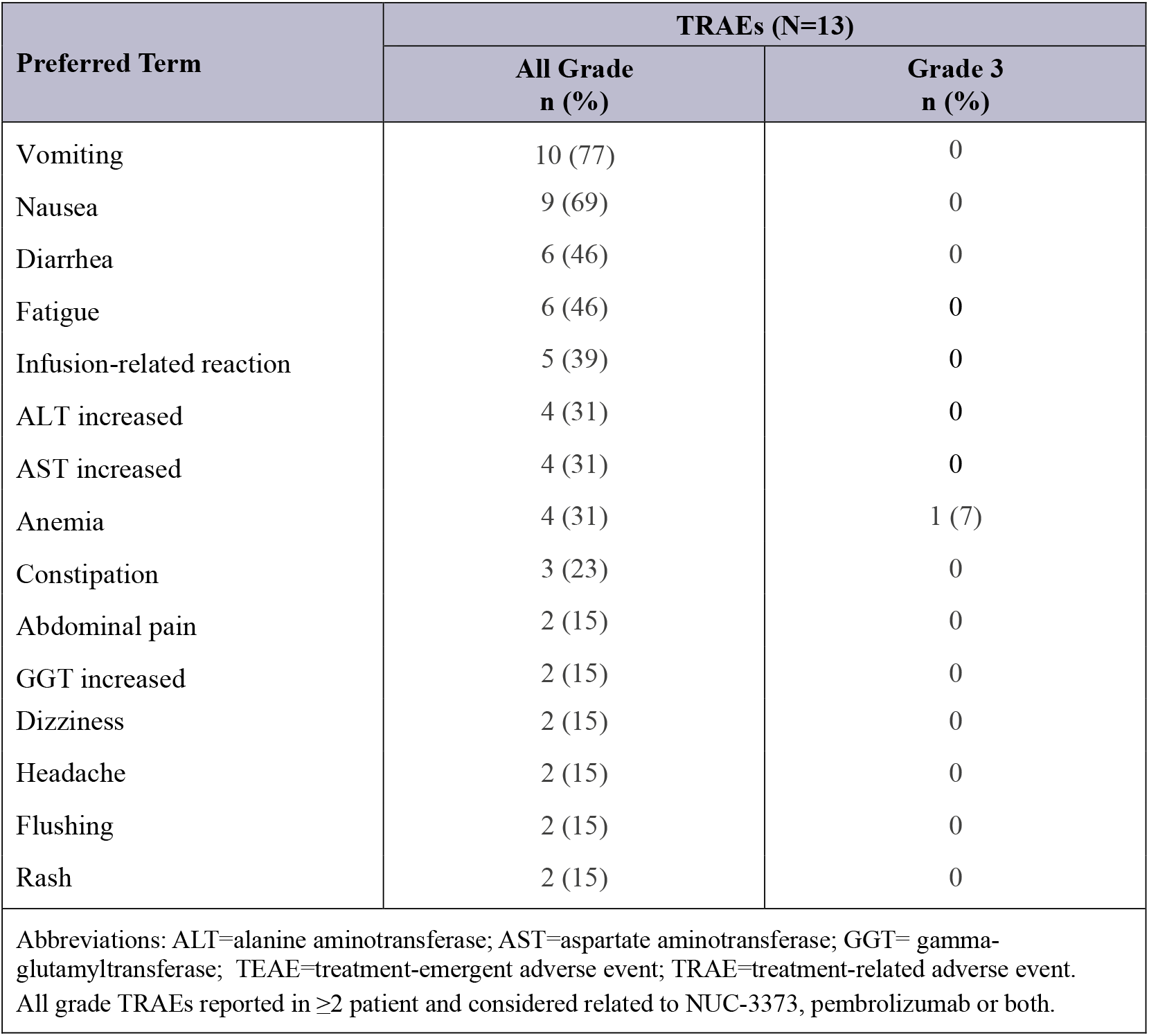
Treatment-related TEAEs in Module 1 (≥15% of patients)

| Preferred Term | TRAEs (N=13) |  |
| --- | --- | --- |
|  | All Grade<br>n (%) | Grade 3<br>n (%) |
| Vomiting | 10 (77) | 0 |
| Nausea | 9 (69) | 0 |
| Diarrhea | 6 (46) | 0 |
| Fatigue | 6 (46) | 0 |
| Infusion-related reaction | 5 (39) | 0 |
| ALT increased | 4 (31) | 0 |
| AST increased | 4 (31) | 0 |
| Anemia | 4 (31) | 1 (7) |
| Constipation | 3 (23) | 0 |
| Abdominal pain | 2 (15) | 0 |
| GGT increased | 2 (15) | 0 |
| Dizziness | 2 (15) | 0 |
| Headache | 2 (15) | 0 |
| Flushing | 2 (15) | 0 |
| Rash | 2 (15) | 0 |
| Abbreviations: ALT=alanine aminotransferase; AST=aspartate aminotransferase; GGT= gamma-glutamyltransferase; TEAE=treatment-emergent adverse event; TRAE=treatment-related adverse event. All grade TRAEs reported in ≥2 patient and considered related to NUC-3373, pembrolizumab or both. |  |  |

A total of 15 patients were enrolled in Module 1, of whom 13 received study treatment (2 patients did not met eligibility requirement at C1D1) . All patients had metastatic disease and the primary tumor types were cutaneous melanoma (n=3), urothelial carcinoma of bladder (n=2), oropharyngeal (n=2), NSCLC (n=2), anal (n=1), sinonasal (n=1), pancreatic (n=1), and pleural mesothelioma (n=1). Patients had received a median of 1 prior line of therapy (range: 0-2) for metastatic disease. Nine patients had received prior immunotherapy treatment, 6 of whom (67%) had disease progression on treatment.

A total of 4 patients were enrolled in Module 2. All patients had metastatic disease and the primary tumor types were NSCLC (n=3) and pleural mesothelioma (n=1). Patients had received a median of 2 prior lines of therapy (range: 1-3) for metastatic disease, and one patient had received prior treatment with paclitaxel.

Further details for individual patients are provided in the Supplementary Data.

### Treatment Exposure

#### Module 1

The DLT evaluation period was 1 cycle in Module 1. No DLTs were observed following treatment of the first 6 patients with 1875 mg/m^2^ NUC-3373 + 400 mg/m^2^ LV + 200 mg pembrolizumab; therefore, the cohort was expanded to 12 patients at the same doses to confirm the recommended dose. Time on treatment was encouraging in this group of heavily pre-treated PD-(L)-1 experienced patients with a variety of different tumor types, with 3 patients remaining on study treatment for prolonged periods of 7, 15, and 19 months (Figure 1). As of 31 August 2025, 1 patient with cutaneous melanoma remains progression-free with a durable partial response (81% reduction, and is receiving a Post-Trial Supply of NUC-3373 + pembrolizumab).

**Figure 1.**
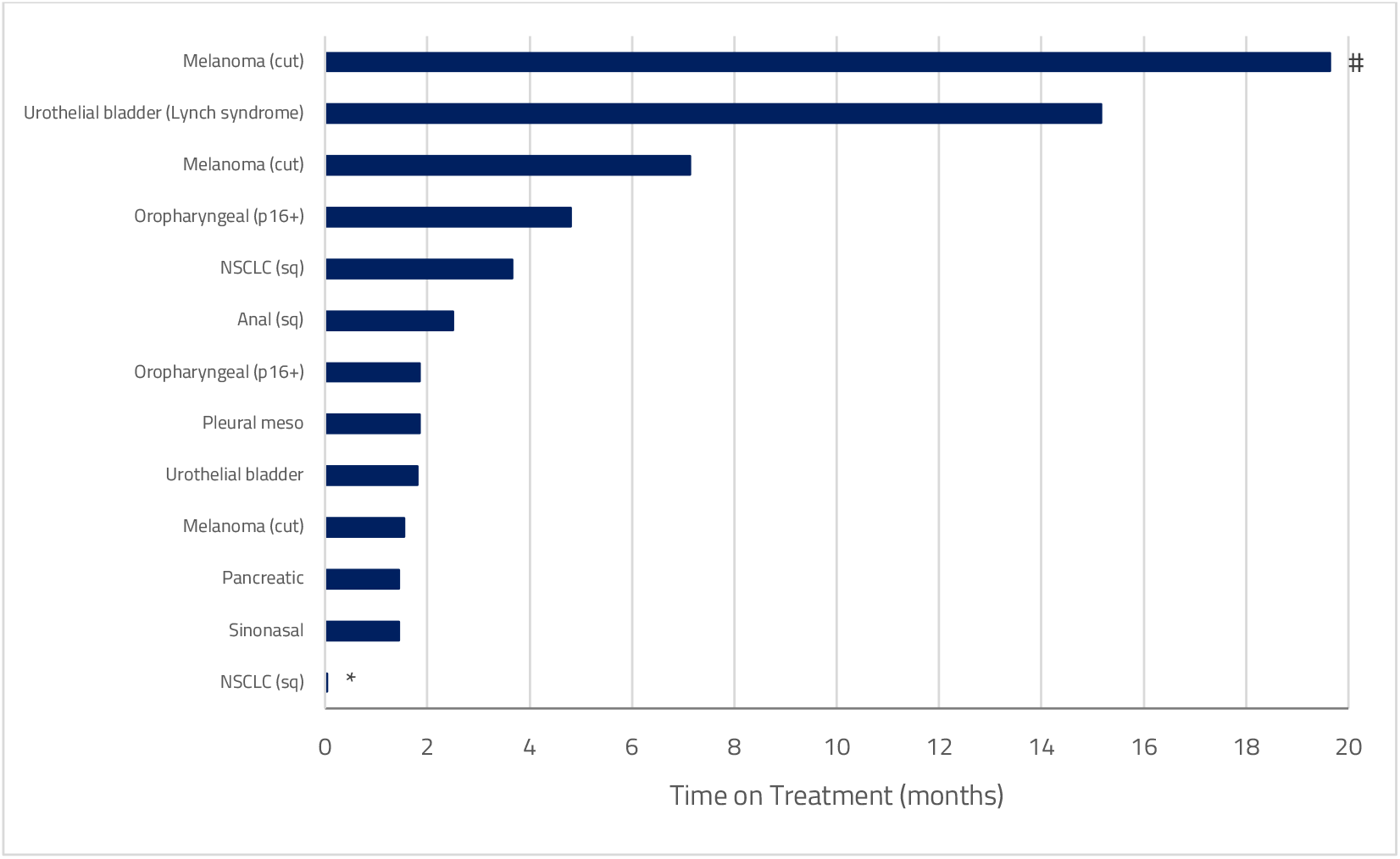
Time on treatment in Module 1. # Data presented as per end of study cut-off date (30 May 2025). This patient was progression-free as of 31 August 2025 (22 months ongoing) * Patient only received 1 dose of study treatment and was not DLT-evaluable

#### Module 2

The DLT evaluation period was 2 cycles in Module 2. No DLTs were observed following treatment of the first 4 patients with 750 mg/m^2^ NUC-3373 + 400 mg/m^2^ LV + 55 mg/m^2^ docetaxel; however, overlapping toxicities of diarrhea, nausea, vomiting, mucositis and fatigue were observed and enrolment to this module was put on hold. Despite this, two patients treated at the first dose level completed more than 6 months of treatment (Figure 2).

**Figure 2.**
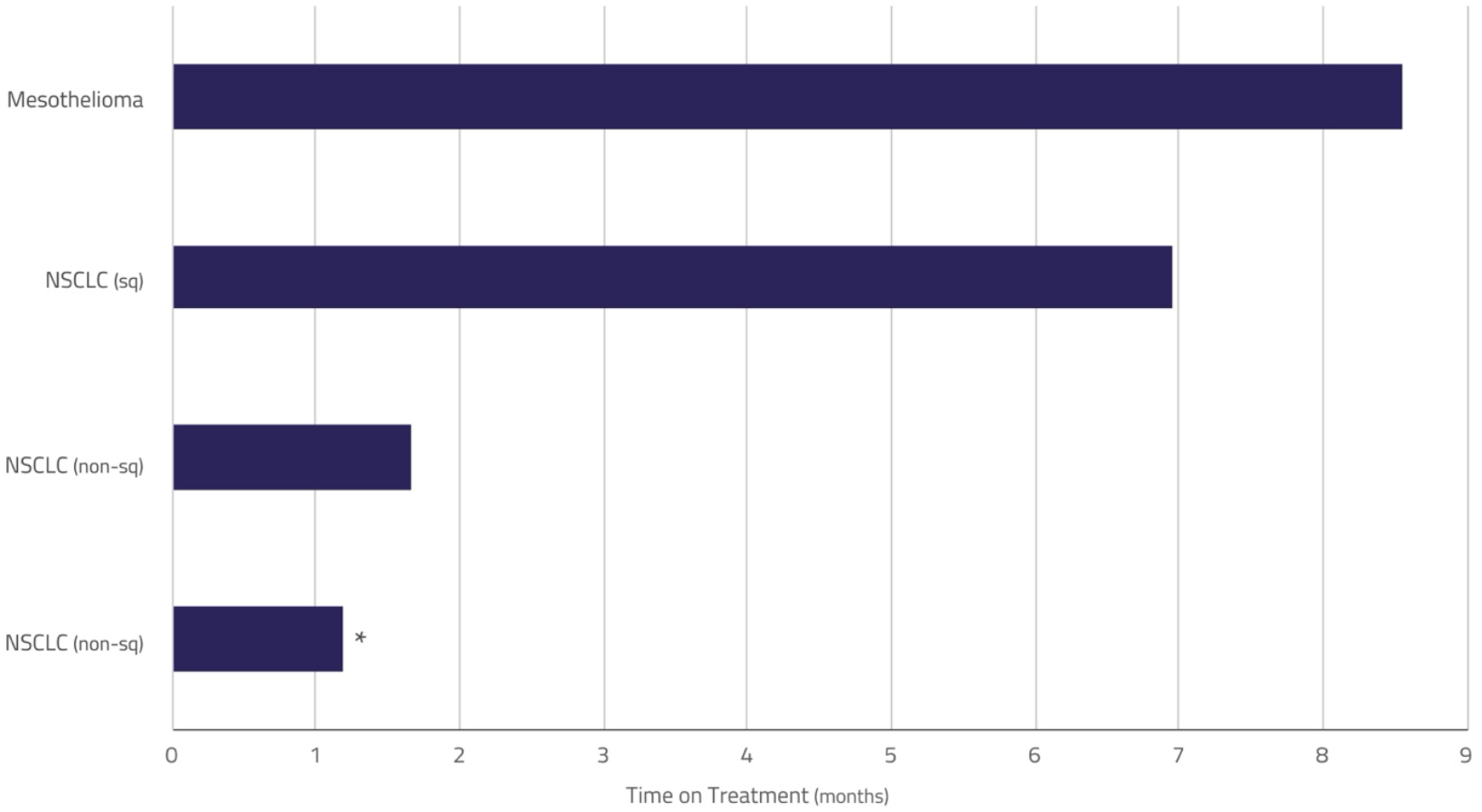
Time on treatment in Module 2. * Patient not DLT-evaluable

Following a Safety Review Committee meeting, the decision was made to stop further enrolment in this module due to tolerability challenges associated with docetaxel. Although no DLTs were observed at this first dose level, the toxicity (largely attributed to docetaxel) was such that it was felt that further dose escalation to single-agent efficacious dose levels of either agent would be difficult. However, given the signs of efficacy of the NUC-3373 + taxane combination, the decision was made to explore other taxanes which may be easier to combine with NUC-3373, rather than proceeding with the NUC-3373 + docetaxel combination.

### Safety

#### Module 1

Thirteen patients were included in the safety population in Module 1. The most common treatment-related TEAEs were vomiting (77%), nausea (69%), diarrhea (46%), fatigue (46%), infusion-related reactions (39%), ALT increased (31%), AST increased (31%), and anemia (31%). The vast majority of treatment-related TEAEs were Grade 1 or 2, with only two patients experiencing Grade 3 TEAEs that were considered to be related to study treatment. One patient had a Grade 3 TEAE of hyponatremia (not listed in table) and one patient had two Grade 3 TEAEs of anemia (all considered to be possibly related to NUC-3373, LV and pembrolizumab). No Grade 4 or 5 treatment-related TEAEs were reported.

#### Module 2

Four patients were included in the safety population in Module 2. The most common treatment-related TEAEs were fatigue (n=4), diarrhea (n=2), hypotension (n=2), nausea (n=2), and stomatitis (n=2). The majority of treatment-related TEAEs were Grade 1 or 2, with three patients experiencing Grade 3 treatment-related TEAEs of fatigue, diarrhea, hypotension, and vomiting (Table 3). No Grade 4 or 5 treatment-related TEAEs were reported.

**Table 3.** Treatment-related TEAEs in Module 2.

| Preferred Term | TRAEs (N=4) |  |
| --- | --- | --- |
|  | All Grade<br>n | Grade 3<br>n |
| Fatigue | 4 | 1 |
| Diarrhea | 2 | 1 |
| Nausea | 2 | 0 |
| Stomatitis | 2 | 0 |
| Constipation | 1 | 0 |
| Vomiting | 1 | 1 |
| Pyrexia | 1 | 0 |
| Anemia | 1 | 0 |
| Decreased appetite | 1 | 0 |
| Hypomagnesaemia | 1 | 0 |
| Hypophosphataemia | 1 | 0 |
| Dizziness | 1 | 0 |
| Dysgeusia | 1 | 0 |
| Alopecia | 1 | 0 |
| Rash | 1 | 0 |
| Hypotension | 1 | 1 |
| Palpitations | 1 | 0 |
| ALT increased | 1 | 0 |
| Rhinorrhea | 1 | 0 |
| Abbreviations: ALT=alanine aminotransferase; TEAE=treatment-emergent adverse event; TRAE=treatment-related adverse event.<br>All grade TRAEs reported in $\geq 1$ patient and considered related to NUC-3373, docetaxel or both. | | |

This module was put on hold due to tolerability challenges with docetaxel at the lowest dose level, as described in the treatment exposure section.

### Efficacy

#### Module 1

Nine patients were evaluable for response. Signals of anti-cancer activity were observed, with durable tumor shrinkages in 2 patients (Figure 3).

**Figure 3.**
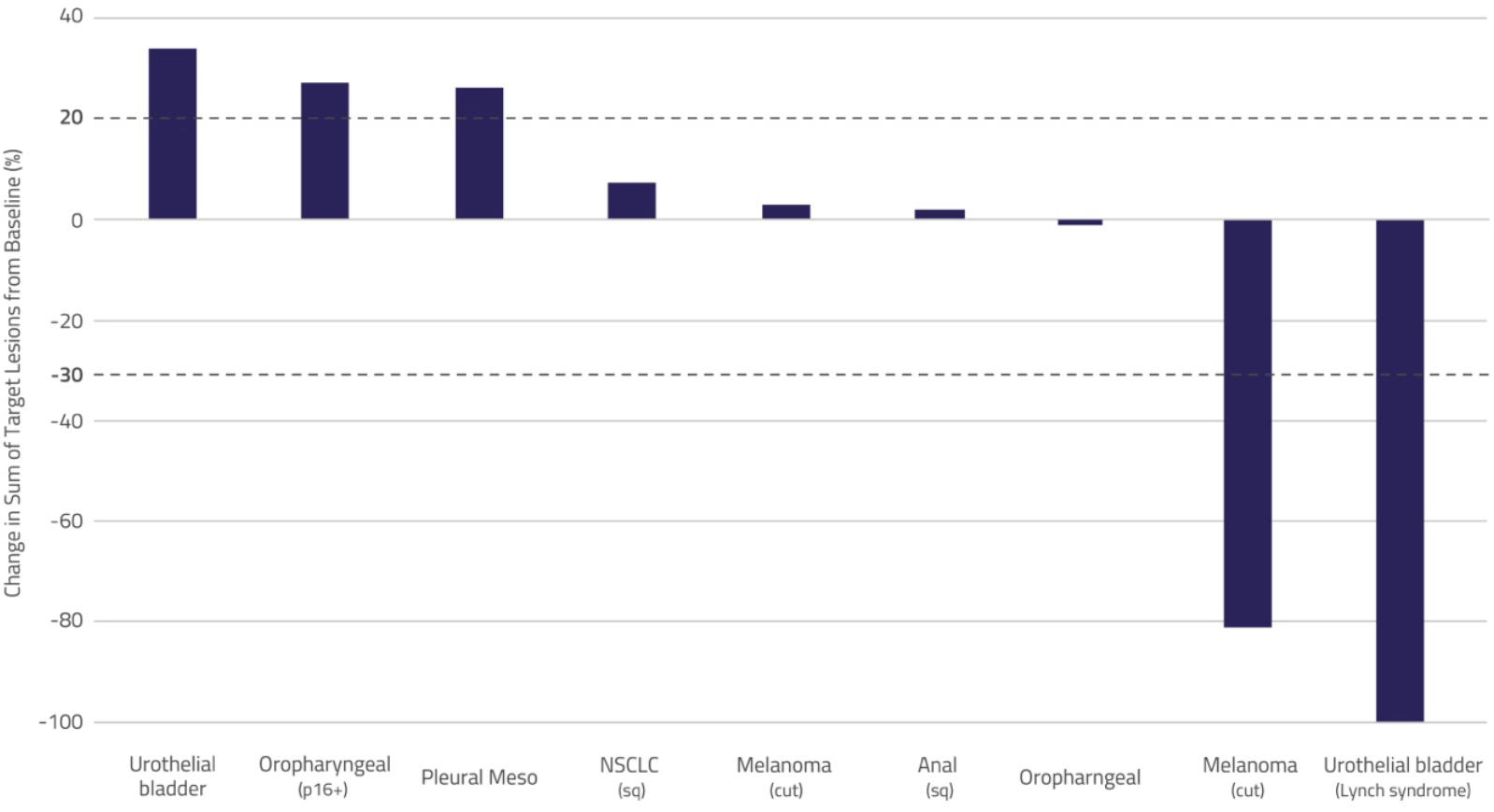
Anti-tumor activity in Module 1.

The best overall response was iPR for 2 patients, iSD for 4 patients, unconfirmed iPD for 2 patients and confirmed iPD for 1 patient, resulting in an objective response rate (ORR) of 22% and a disease control rate (DCR) of 67%. One patient had a 100% reduction in the sum of target lesions, but non-target lesions were present and so was not considered a complete response (iCR). Another patient had a confirmed partial response (iPR) with an 81% reduction in tumor volume.

Further information on the two patients with PRs is presented in the case studies below.

A patient with urothelial carcinoma of bladder (Lynch syndrome) had received 2 prior lines of therapy. They received gemcitabine plus cisplatin in the adjuvant setting (discontinued after 2 months due to myelosuppression) and atezolizumab in the metastatic setting (best response of PR and discontinued after 23 months as had received maximum allowable therapy). They had 1 target lesion in the lung. The patient received treatment with 1875 mg/m^2^ NUC-3373 plus 200 mg pembrolizumab and had a 100% reduction in the target lesion; however, non-target lesions were present so this response was considered a confirmed iPR and not a iCR. The patient remained on treatment for approximately 15 months before discontinuing due to progressive disease.

A patient with cutaneous melanoma had received 2 prior lines of therapy. They received pembrolizumab (best response of PD within 5 months) and trametinib plus dabrafenib (discontinued trametinib after 1 month due to toxicity and achieved best response of SD on dabrafenib before experiencing PD after 7 years). They had 1 target lesion in the lymph nodes (bilateral). The patient received treatment with 1875 mg/m^2^ NUC-3373 plus 200 mg pembrolizumab and had an 81% reduction in tumor volume (confirmed iPR). The patient remained on treatment in the study for approximately 19 months before receiving post-trial NUC-3373 + pembrolizumab. This patient remains progression-free with a durable partial response at 22 months as of 31 August 2025.

#### Module 2

Three patients were evaluable for response. The best overall response was SD for 2 patients and PD for 1 patient. Further information on the two patients with SD is presented in the case studies below.

A patient with pleural mesothelioma had received 3 prior lines of therapy. They received cisplatin plus pemetrexed (PD within 4 months), nivolumab (PD within 4 months), and carboplatin plus pemetrexed (PD within 1 month). This patient had progressive disease and a high tumor burden at study entry. They had 4 target lesions (2x lymph node and 2x mediastinum). The patient received treatment with 750 mg/m^2^ NUC-3373 plus 55 mg/m^2^ docetaxel and achieved prolonged SD of more than 18 months. The patient received 4 cycles of NUC-3373 plus docetaxel followed by 5 cycles of NUC-3373 alone before coming off treatment due to patient choice (treatment fatigue). The patient has not experienced PD and the Investigator believes the patient derived clinical benefit (durable disease control) from NUC-3373.

A patient with NSCLC (squamous) had received 2 prior lines of therapy. They received carboplatin plus paclitaxel plus pembrolizumab (SD for 2 months) and pembrolizumab as maintenance (PD within 21 months). They had 1 target lesion in the lung. The patient received treatment with 750 mg/m^2^ NUC-3373 plus 55 mg/m^2^ docetaxel and achieved prolonged SD of 7 months. The patient received 6 cycles of NUC-3373 plus docetaxel followed by 2 cycles of NUC-3373 alone before coming off treatment due to disease progression.

## Discussion

This Phase Ib/II modular study was designed to evaluate optimal combination partners for NUC-3373 in patients with advanced solid tumors. Two modules were opened and here we present data from the Phase Ib part of Modules 1 and 2. In Module 1, NUC-3373 was evaluated in combination with LV and pembrolizumab for the treatment of patients with advanced solid tumors. In Module 2, NUC-3373 was evaluated in combination with LV and docetaxel for the treatment of patients with NSCLC or pleural mesothelioma.

Results from Module 1 show signals of anti-cancer activity, including meaningful clinical benefit in some patients, and tolerability, providing support for further exploration of NUC-3373 plus pembrolizumab in future studies. These early clinical results build on nonclinical studies and provide further support that NUC-3373 may promote an anti-tumor immune response and potentiate the activity of immune checkpoint inhibitors. In previous *in vitro* studies it has been shown that NUC-3373 binds to TS which accumulates in the cytoplasm and leads to release of DAMPs [9, 10], which were predicted to increase the likelihood of an anti-tumor inflammatory response.

Results from Module 2 suggest that, although no DLTs were observed at the first dose level, the toxicity (largely attributed to docetaxel) was such that further dose escalation to single-agent efficacious dose levels of either agent would have been difficult. Given that the docetaxel starting dose (55 mg/m^2^) was lower than the standard dose used in NSCLC (75 mg/m^2^), dose de-escalation of docetaxel was not considered a viable option. Docetaxel is the current standard of care for NSCLC patients without targetable alterations who progress on PD-(L)1 inhibitor-based therapy; however, it is associated with modest clinical benefit (median progression-free survival of 3-4 months) and substantial toxicity [21]. Achieving disease stabilization in hard-to-treat populations such as NSCLC and mesothelioma further supports the hypothesis that NUC-3373 may be an attractive treatment option for primary lung malignancies based on nonclinical studies showing that NUC-3373 is a potent TS inhibitor.

Overall, data from NuTide:303 suggest that NUC-3373 may be an effective combination partner for pembrolizumab in a variety of solid tumors. Although the combination of NUC-3373 and docetaxel may not be tolerable, different NUC-3373 regimens with an alternative taxane could be a promising treatment option in lung cancer and are under consideration.

## Data Availability

All data produced in the present study are contained in the manuscript and supplementary data

