## Supplemental data for "A Phase Ib/II multi-arm, dose finding and expansion study of a novel thymidylate synthase inhibitor with immune modulating properties, NUC-3373, in combination with pembrolizumab or docetaxel in patients with advanced solid tumors (NuTide:303)"

1) Birmingham NHS Foundation Trust/University of Birmingham, UK
2) King's College London, Guy's Hospital, London, UK
3) The Christie NHS Foundation Trust
4) Beatson West of Scotland Cancer Centre/University of Glasgow, UK
5) University of St Andrews, UK
6) NuCana plc
7) University of Manchester, UK

**Disclosure:** GM has performed consulting and advisory roles at Roche, Boehringer Ingelheim, Mina therapeutics, D2G, BMS, MSD, and AstraZeneca; has participated in speakers' bureaus for Roche, BMS, MSD, AstraZeneca, Servier, and Bayer; and has received research funding from BMS, MSD, AstraZeneca, and Plexxikon. JS owns stock in Epsilogen Ltd (co-founder) and Avacta Ltd; holds board membership at Apobec Discovery Ltd; has performed advisory roles at Apobec Discovery Ltd, Avacta Ltd, AstraZeneca, BioNTech, BMS and Roche; has received assistance with attendance at international meetings from Starpharma and MSD; and has received reimbursement for recruitment to clinical trials from Achilles, Gilead, GSK, IO Biotech, MSD, Roche, RS Oncology, and Starpharma. RW has performed consultancy and advisory roles at Alcimed, Amphista Therapeutics, BMS, Boehringer Ingelheim, CV6 Therapeutics, NuCana, RIN Institute and Takeda; has received assistance with attendance at international meetings from NuCana and Takeda; and has received honoraria for educational meetings from Bayer, Merckgroup, Pierre Fabrier, Servier and Takeda. DJH is a part-time employee of NuCana. EO and JB are full-time employees of NuCana. FT has performed consulting and advisory roles at CytomX, Kite, Gilead, Scenic Biotech, Immatics, T-Knife Therapeutics, F-Star, Grey Wolf Therapeutics, and AstraZeneca.

### Supplementary Data

#### Table of Contents

*Definition of DLT*........................................................................................4
*Summary of Patients*.....................................................................................5

### Definition of DLT

DLT was defined as any of the following occurring during Cycle 1 (Module 1) or Cycles 1 and 2 (Module 2) that were not due to the protocol disease indication or to a known concurrent medical condition and were judged as clinically significant and related to study treatment.

| **For dose escalation, DLTs were defined as follows** |
|---|
| **Any Grade 4 AEs, with the exception of:** |
| Neutropenia lasting <72 hours that is not associated with fever or other clinical symptoms |
| Lymphopenia |
| Electrolyte abnormalities that are not associated with clinical sequelae and are corrected with appropriate management or supplementation within 72 hours of the onset |
| **Any Grade 3 AEs, with the exception of:** |
| Nausea and vomiting persisting for <2 days after optimal anti-emetic therapy |
| Thrombocytopenia without significant bleeding (including platelet transfusions) |
| Diarrhea persisting for <2 days after optimal anti-diarrheal treatment |
| Hypertension persisting <7 days after treatment |
| Infection or fever in the absence of neutropenia persisting <5 days |
| Rash or photosensitivity persisting <7 days after treatment |
| Fatigue persisting <7 days |
| Liver function test elevations (AST/ALT) persisting <7 days after treatment with corticosteroids |
| Immune-related adverse events persisting <7 days after treatment with corticosteroids<br>Alopecia |
| **The following Grade 2 AEs were to be adjudicated as DLTs:** |
| Total bilirubin CTCAE v5 ≥Grade 2 with simultaneous ALT/AST CTCAE v5 ≥Grade 2 |
| Pneumonitis persisting >7 days despite treatment with corticosteroids |
| Eye pain or reduction of visual acuity that does not respond to topical therapy and does not improve to Grade 1 severity within 2 weeks of the initiation of topical therapy OR requires systemic treatment |
| Other clinically significant toxicities (cardiotoxicity, neurotoxicity) including a single event or multiple occurrences of the same event that lead to a dosing delay of >7 days in Cycle 1 |

### Summary of Patients

| Module 1 | | |
|---|---|---|
| **Disease** | **Prior Treatment for Metastatic Disease** | **Metastatic Sites** |
| NSCLC<br>(squamous) | 1. docetaxel | 1 x lung |
| Sinonasal Carcinoma | 0 | 1 x nose |
| Pleural Mesothelioma | 1. ipilimumab + nivolumab | 2 x pleura |
| Urothelial Bladder | 1. gemcitabine + cisplatin<br>2. avelumab | 2 x liver |
| Oropharyngeal<br>(p16+) | 1. nivolumab<br>2. investigational agent + atezolizumab | 2 x lung<br>1 x mediastinum<br>2 x pelvis |
| Oropharyngeal | 1. nivolumab | 2 x lung<br>1 x mediastinum |
| Urothelial Bladder<br>(Lynch Syndrome) | 1. atezolizumab | 1 x lung |
| Cutaneous Melanoma | 1. pembrolizumab<br>2. dabrafenib + trametinib | 1 x lymph node |
| Cutaneous Melanoma | 1. ipilimumab + nivolumab | 1 x lung<br>1 x liver<br>1 x mediastinum |
| Pancreatic Adenocarcinoma | 1. FOLFIRINOX<br>2. gemcitabine + cisplatin | 2 x liver |
| Anal<br>(squamous) | 1. paclitaxel + carboplatin<br>2. capecitabine + mitomycin c | 2 x lung<br>1 x lymph node |
| NSCLC<br>(squamous) | 1. pembrolizumab | 2 x lymph node<br>2 x lung |
| Cutaneous Melanoma | 1. ipilimumab + nivolumab | 1 x lymph node |

| Module 2 | | |
|---|---|---|
| **Disease** | **Prior Treatment for Metastatic Disease** | **Metastatic Sites** |
| NSCLC<br>(squamous) | 1. carboplatin + paclitaxel + pembrolizumab | 1 x lung |
| NSCLC<br>(non-squamous) | 1. pembrolizumab<br>2. carboplatin + pemetrexed | 2 x non-lymph node |
| Pleural Mesothelioma | 1. cisplatin + pemetrexed<br>2. nivolumab<br>3. carboplatin + pemetrexed | 2 x lymph node<br>2 x mediastinum |
| NSCLC<br>(non-squamous) | 1. gemcitabine + cisplatin<br>2. atezolizumab | 1 x lung<br>2 x lymph node<br>1 x muscle/soft tissue |
